# Comparing Decision Tree-Based Ensemble Machine Learning Models for COVID-19 Death Probability Profiling

**DOI:** 10.1101/2020.12.06.20244756

**Authors:** Carlos Pedro Gonçalves, José Rouco

## Abstract

We compare the performance of major decision tree-based ensemble machine learning models on the task of COVID-19 death probability prediction, conditional on three risk factors: *age group, sex* and *underlying comorbidity or disease*, using the US Centers for Disease Control and Prevention (CDC)’s COVID-19 case surveillance dataset. To evaluate the impact of the three risk factors on COVID-19 death probability, we extract and analyze the conditional probability profile produced by the best performer. The results show the presence of an exponential rise in death probability from COVID-19 with the age group, with males exhibiting a higher exponential growth rate than females, an effect that is stronger when an underlying comorbidity or disease is present, which also acts as an accelerator of COVID-19 death probability rise for both male and female subjects. The results are discussed in connection to healthcare and epidemiological concerns and in the degree to which they reinforce findings coming from other studies on COVID-19.

## 1. Introduction

The coronavirus disease 2019 (COVID-19), caused by severe acute respiratory syndrome coronavirus 2 (SARS-CoV-2), has become a major global health problem. In the context of the SARS-CoV-2 pandemic, the development of data-driven death risk profiling is, therefore, a relevant matter, providing healthcare authorities with results that can be used for policy-making and in the planning of a response to the COVID-19 crisis, also, from an epidemiological standpoint, a death risk profile provides for a relevant characterization of the virus and the corresponding disease, conditioned on risk factors.

Different studies have identified *age group, sex* and *underlying comorbidity or disease* as critical factors for disease severity and mortality risk [1-5]. In the current work, we use these three factors as feature variables in machine learning models for extracting a profile for COVID-19 death probability, conditional on these three risk factors, using the US Centers for Disease Control and Prevention (CDC)’s COVID-19 case surveillance data sample^1^ which contains a large sample of COVID-19 confirmed cases with records for these three factors.

In regards to the *age group* variable, the CDC defined an ordinal scale of nine age groups, we use this ordinal feature variable along with the qualitative variables *sex* and *underlying comorbidity or disease*^*2*^. Together, these feature variables provide a set of 36 alternative configurations for the three critical factors, *age group, sex* and having an *underlying comorbidity or disease*. Using these three feature variables, we compare the performance of major tree-based ensemble machine learning models with that of standard logistic regression (used as a baseline) in the prediction of COVID-19 death probability, and select the alternative that shows the best reliability performance in order to extract and study a death probability profile conditional on the combinations of values of these three feature variables.

Tree-based ensemble machine learning models have an advantage when working with ordinal and nominal variables defined on different scales, since they can work directly with qualitative features, not suffering from possible problems resulting from performing arithmetic operations on different qualitative variables or assuming distance metrics over these types of variables. The fact that we are dealing with only qualitative features, since age comes pre-classified by the CDC into the *age group* ordinal variable and the remaining two feature variables are binary and qualitative, this makes tree-based ensemble models more sound on a statistical analysis and interpretability basis, a point to which we will return in the *materials and methods* section.

In the medical and epidemiological context, tree-based ensemble algorithms have been successfully applied for mortality risk prediction, both in terms of death event prediction and mortality risk scoring [6-10]. In the current work, we will be comparing the performance of major tree-based ensemble models, namely: random forests, extremely randomized trees with and without bootstrap sampling, AdaBoost, boosted random forests, boosted extremely randomized trees with and without bootstrap sampling, gradient boosted decision trees and LighGBM-based histogram boosting trees.

To compare the performance of the different models, since we are dealing with probability prediction rather than a standard classification problem, main classification metrics such as the area under receiver operating characteristic’s (ROC) curve (AUC) are no longer appropriate measures [11], in this case, reliability plots and Brier loss are usually used for performance evaluation [11-15].

The work is divided into three sections: **materials and methods** (section 2), **results** (section 3) and **discussion** (section 4). In the **materials and methods** section, we provide for an exploratory analysis of the three factors in the database, also providing for an inferential analysis of death proportions for each risk factor.

After the exploratory analysis, we review the main probability prediction performance evaluation methods and the main decision tree-based ensemble algorithms that we will be testing.

In the **results** section, we apply and compare the performance of the different machine learning models, selecting the best performing alternative in order to extract a death probability profile that we analyze in the second part of the **results** section. In the **discussion** section, we address the main implications of the resulting profile for the fight against the SARS-CoV-2 pandemic.

## 2. Materials and Methods

### 2.1. The Dataset and Exploratory Data Analysis

The CDC’s COVID-19 case surveillance data is available for public use and, in the November 4 updated version, which we use in the present work, contains a sample of 5,760,066 individuals, with 5,462,778 confirmed cases and 297,288 probable cases. Since our goal is to study the death probability of people with confirmed COVID-19, conditioned on the three major risk factors that we are addressing: *age group, sex* and the presence of *underlying comorbidity or disease*.

Our focus is on the confirmed cases and in these three feature variables that are present in the database. One of the main reasons to choose this database was the fact it contains these three risk factors for a large number of individuals.

Of the 5,462,778 confirmed cases, there are missing values for all of the three risk factor variables that we work with, these missing values must be removed in a preprocessing stage in order to be able to work with the machine learning models in a way that may produce a death probability profile given definite values for the three critical risk factors, which is our main objective.

For the sex variable, there are three defined sex categories, *male, female* and *other*, which might lead to the possibility of working with the *sex* variable as a non-binary feature variable, in this case, it is not clear if the third category *other* refers to gender identification rather than biological sex, in which case, it would correspond to a third gender, the problem here is that there is an insufficient number of cases in the classification, which is 14 for the category *other*, while the male sample size is 391,060 and the female is 436,541. The small sample size of the category *other* does not provide a sufficient number of cases for machine learning, these 14 cases are all laboratory confirmed cases and neither of them died.

Removing all the missing values from the features plus extracting the 14 cases classified as *other* in the sex feature variable, we get a sample of 827,601 confirmed cases of COVID-19, with definite values for the target and feature variables. In this sample, 66,686 individuals have died while 760,915 have not, which gives a sample death rate of around 8.7639% for the confirmed cases, in the database.

In **table 1**, we provide the sample distributions for the final database of 827,601 confirmed cases and each of the three critical factors. In terms of medical condition, the majority of cases in the database (430,758) exhibit an underlying comorbidity or disease. Regarding the sex variable, there are more female cases than male (436,541 female cases against 391,060 male cases), of the female cases, 52.4283% show an underlying comorbidity or disease while, for the males, this percentage is 51.6256%.

**Table 1:** Sample distribution by feature variable.

| Features |  | Number of Cases |
| --- | --- | --- |
| **Medical condition** | Yes | 430,758 |
|  | No | 396,843 |
| **Sex** | Male | 391,060 |
|  | Female | 436,541 |
| **Age** | 0 - 9 Years | 23,564 |
|  | 10 - 19 Years | 74,291 |
|  | 20 - 29 Years | 149,286 |
|  | 30 - 39 Years | 121,476 |
|  | 40 - 49 Years | 118,988 |
|  | 50 - 59 Years | 124,876 |
|  | 60 - 69 Years | 95,906 |
|  | 70 - 79 Years | 61,150 |
|  | 80+ Years | 58,064 |

Regarding the nine age group categories, the main frequency counts are situated in the four age groups from 20-29 years to 50-59 years, all with more than 100,000 cases.

The presence of underlying comorbidity or disease rises with age with a pattern that is close to a sigmoid curve, as shown in **figure 1**.

**Fig. 1:**
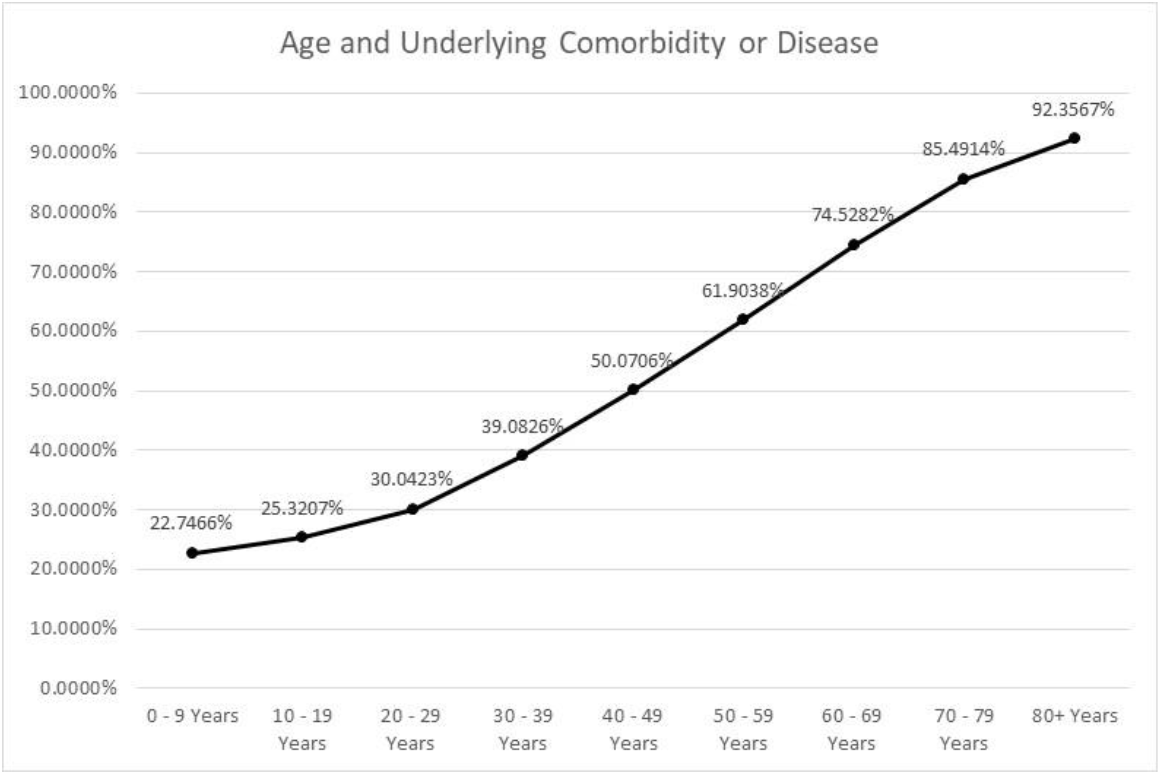
Percentage of cases with underlying comorbidity or disease by age group.

In **table 2**, we show the 95% confidence intervals for the death proportion conditional on each feature variable. The results show that having an underlying comorbidity or disease significantly rises the death risk. The male sex also indicates a higher level of death risk, in inferential terms. The death risk also rises with age, with the age group of 80+ Years exhibiting a 95% confidence interval for the death proportion with a lower bound of 0.505483 and an upper bound of 0.513633.

**Table 2:** 95% confidence intervals for the death proportion conditional on each feature variable.

| Features |  | 95% Confidence Intervals for Death Proportion |
| --- | --- | --- |
| Medical condition | Yes | (0.147151, 0.149275) |
|  | No | (0.006904, 0.007431) |
| Sex | Male | (0.092362, 0.094188) |
|  | Female | (0.068454, 0.069962) |
| Age | 0 - 9 Years | (0.000519, 0.001311) |
|  | 10 - 19 Years | (0.000373, 0.000718) |
|  | 20 - 29 Years | (0.001555, 0.001988) |
|  | 30 - 39 Years | (0.006051, 0.006963) |
|  | 40 - 49 Years | (0.015765, 0.017220) |
|  | 50 - 59 Years | (0.042469, 0.044742) |
|  | 60 - 69 Years | (0.119847, 0.123998) |
|  | 70 - 79 Years | (0.272681, 0.279785) |
|  | 80+ Years | (0.505483, 0.513633) |

The death proportion’s 95% confidence intervals’ lower and upper bounds exhibit an exponential rise with age, as shown in **figure 2**, which indicates, inferentially, with 95% confidence, an exponentially rising death probability for confirmed COVID-19 cases. We will see that the machine learning models capture well this exponential profile.

**Fig. 2:**
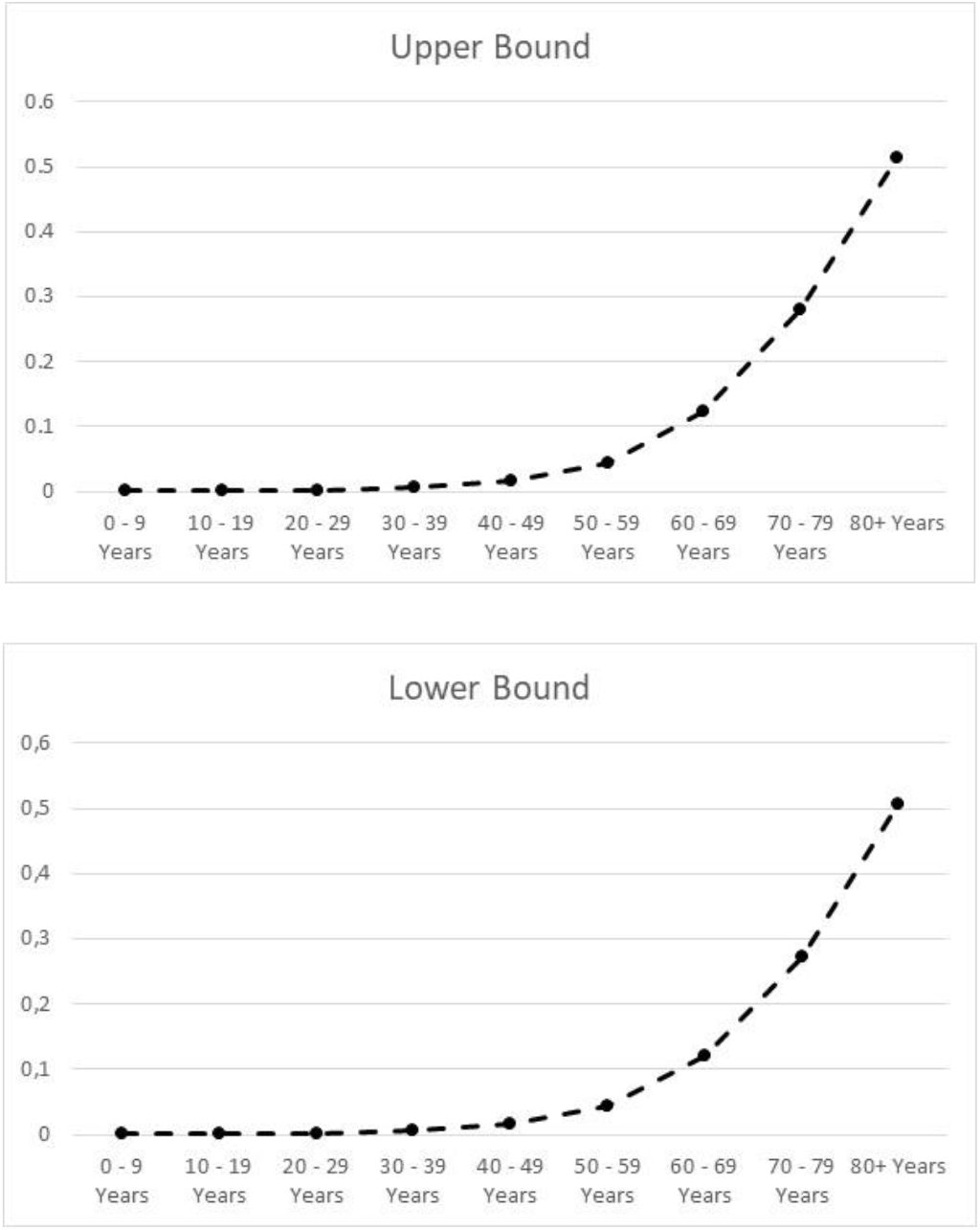
Upper and lower bounds of 95% confidence intervals for the death proportion conditional on the age group variable.

Considering the feature space, as stated before, we have a finite discrete qualitative feature set, which is comprised of 36 alternative feature values, this type of structure has consequences in the evaluation methodologies for machine learning models applied to probability prediction, as we now discuss.

### 2.2. Evaluation Methods of Machine Learning Algorithms Applied to Probability Prediction Problems

Probability prediction problems, in the context of supervised learning, involve the prediction of class probabilities rather than class labels, therefore, the main objective is to predict probabilities in such a way that the distribution predicted by the learning algorithm fits well the sample proportions [11-15]. In medical applications, the risk profiling associated with the development of a disease and mortality risk assessment fits into this type of problem. When dealing with an epidemiological scenario, the profiling of death probabilities, conditional on subjects’ features, can help identify risk groups and guide response strategies. In this context, when dealing with the assessment of death probability profiles, the target is binary, with 0 labeling a person who survived and 1 a person who died^3^.

When evaluating different machine learning algorithms in probability profiling, for a binary target, the performance metrics must be directed at calibration performance [11], in this sense, while the AUC can be reported and used as a class label prediction metric, in probability profiling tasks, it does not provide for a good indicator of performance [11]. In this case, evaluation methods have been proposed and used, with the Brier loss and reliability plot-based metrics standing out for these tasks [11-15].

The Brier loss, also known as Brier score, for a binary target and a sample of size *N* is calculated as the mean squared difference between the predicted Bernoulli success probabilities 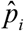, produced by the machine learning model for each case, and the observed sample values *y*_*i*_ [11-13]:

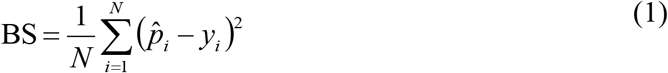

The lower the Brier loss is, the better is the performance of the machine learning model at predicting the probabilities. Besides the Brier loss, which works with the squared deviations between the predicted probabilities for each sample case and the corresponding true labels, reliability plot-based metrics can be calculated for the direct comparison between observed proportions and predicted probabilities [14,15]. Reliability plots are major tools to visually evaluate probability prediction calibration and have been used to detect risk assessment biases in measures like the Brier loss, as analyzed in [13] about the use of this score for medical diagnosis. The advantage of reliability plots is that they address directly probability prediction, since one is comparing predicted probabilities with observed frequencies. However, when the feature variables range in a continuous scale, binning must be used, with the researcher selecting the number of bins for the plot, this has led to the calculation of approximations to reliability plot error metrics, estimated from binning [14,15].

On the other hand, when feature variables are qualitative, which is our case, then, no additional binning is needed, in these cases, an exact feature space can be used to calculate the Bernoulli success proportions, so that the reliability plot works with a frequencist underlying logic, since it compares, for each alternative feature variables’ configuration, observed Bernoulli success proportions and the machine learning-based probability predictions, without any binning involved. If the number of samples for each qualitative feature variables’ configuration is large enough for the law of large numbers to hold, then, we can use the reliability plot’s underlying frequencist logic and estimate calibration metrics, from the reliability plot, without any binning decisions involved in the process.

In our case, reliability-plot main metrics can be employed to evaluate the machine learning models’ performance, without binning impact considerations entering into play, since, as shown in the previous subsection, we are dealing with a finite discrete set of feature variables and an exact conditional sample proportions’ distribution, without having to resort to any additional binning to get the reliability plot^4^.

Indeed, when dealing with a finite discrete set *X* of alternative vector values for feature variables, which is the case when only qualitative features are being used, the reliability plot becomes the plot of the following ordered pairs’ finite set 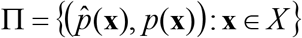, where 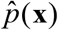 is the predicted conditional probability for the Bernoulli success by the machine learning model when an individual is characterized by the feature vector **x** and *p*(**x**) is the corresponding conditional sample proportion.

In this case, of finite discrete alternative feature values’ set *X*, the number of points *N*_*r*_ in the reliability plot coincides with the size of the set *X*, that is, it coincides with the size of the feature space, therefore, working with the reliability pairs, one can calculate, for a such reliability plot, a *Reliability Plot Root Mean Square Error* (RPRMSE) defined as follows:

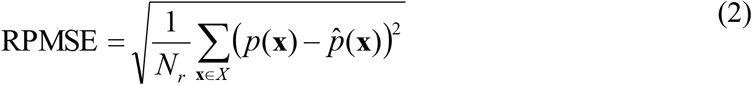

If the number of calibration points *N*_*r*_ is sufficiently large, the reliability plot’s RPRMSE becomes a statistically relevant metric for the deviation between the machine learning-predicted success probabilities and the observed success proportions.

The lower the RPRMSE is, the closer are the predicted probabilities to the observed sample proportions. This measure has the advantage that it directly addresses the reliability plot and penalizes deviations where risk is underestimated or overestimated, which is a problem that has been identified in regards to the Brier loss in the medical context, since this loss can penalize less cases of risk underestimation as addressed in [13]. The reliability plot’s explained variance score is another possible metric that can also be estimated for a reliability plot with a finite discrete feature space, when the number of plotted reliability points is sufficiently large (that is, when *N*_*r*_ is large), so that we have the following reliability plot explained variance (RPEV):

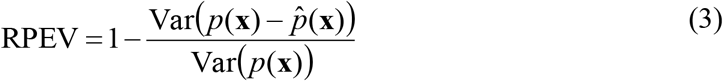

These metrics, which are used in machine learning regression problems, can be applied to the reliability plot, worked as a scatter plot, where the observed Bernoulli success proportions are compared with the predicted success probabilities.

Reliability plot-based metrics have the advantage that they directly address the probability prediction task, comparing observed proportions with predicted probabilities. A basic statistical assumption is that there is a sufficiently large number of samples for each alternative feature variables’ configuration so that the law of large numbers applies to each estimated sample proportion, in this case, the closest the predicted probabilities are to the sample proportions, with the law of large numbers holding, the better performing is a machine learning model at probability prediction, in the sense that it may be capturing better the theoretical probabilities, this is a basic assumption underlying the reliability plot itself, which compares observed proportions with predicted probabilities.

For model selection, given the critical role of reliability plots in finding well-calibrated models that neither underestimate nor overestimate risk, overcoming some of the reported problems of the Brier loss applied to the medical context [13], and considering also the nature of the feature variables space that we are working with, we will use the RPRMSE and the RPEV as main model selection criteria, even though we also report both the Brier loss and the AUC metrics.

### 2.3 Tree-based Ensemble Machine Learning Models

Decision trees are discriminant-based hierarchical models for supervised learning that lead to a bridge with rules-based artificial intelligence systems, since the resulting final tree can be expressed as a set of branching *if-then rules*, which makes these models interpretable and good for use in clinical diagnosis, another advantage of tree models is that for ordinal or nominal feature variables, a numeric encoding is less problematic than for estimators that work with sums of values of the feature variables multiplied by weights, as is the case with logistic regression and also for other models like neural networks, in these last cases, if not carefully done, the encoding can lead to problems when weighted sums over numerically encoded qualitative feature variables are used to produce a prediction as well as when different numerical encoding scales are warranted.

The downside of decision trees is, however, that single-tree models suffer from high variance and sometimes can lead to poorer results when compared with alternative machine learning models, which reduces their applicability. Tree-based ensemble models were developed to overcome the problem of the high variance and lower performance, making tree-based methods competitive as machine learning solutions.

There are two major tree-based ensemble methods: randomization and boosting [16]. Randomization methods try reduce the variance of trees by introducing randomization into the learning algorithm and/or into the learning sampling process, with the predictions being aggregated by an average or a majority vote, boosting methods use weak learners and an adaptive process for increasing performance. Randomization and boosting can also be combined with each other. The randomization models that we will compare are the following:

- Random forest with bootstrap sampling and out-of-bag (OOB) scoring;
- Extremely randomized trees without bootstrap sampling;
- Extremely randomized trees with bootstrap sampling and OOB scoring.

Random forests were introduced by Breiman [17] as a way to improve tree-based ensembles’ performance, while not increasing the bias significantly [16,17]. Random forests have been applied in the medical context [7-10], including, most recently, to the SARS-CoV-2 pandemic [18,19].

In a random forest, each tree in the ensemble is sampled independently, with the same distribution for all the trees in the forest, and such that the generalization error converges almost surely to a limit with an increase in the number of trees [17]. The method employs bootstrap samples, where each tree in the ensemble is trained on a sample drawn with replacement from the training set, which makes random forests effective learners for smaller datasets, this bootstrap sampling can be enhanced with *out-of-bag* (OOB) samples, used to estimate the generalization accuracy, which may reduce the tendency of trees to overfit and increase the performance on test data. For small data, where training, validation and test samples would greatly reduce the number of examples, the use of OOB samples is a useful way to integrate a validation process in learning, increasing the generalization ability and reducing overfitting.

Another source of randomness in random forest models comes from the criterion for splitting nodes in the trees, in this case, the best fit can be found either from all input features, or, alternatively, by taking a random subset of features of a predefined size. The bootstrap sampling with OOB scoring and the random choice of features for each split are aimed at decreasing the variance and reducing the tendency for overfitting.

The variance reduction and increased performance is strongly linked to the randomness in the ensemble, which tends to lead to trees with low correlated or even decoupled prediction errors, so that by taking the average over the trees, the overall error and variance tend to be reduced. Throughout the work, we will use Python’s *scikit-learn* library, which introduces a further element over the original random forest model proposed by Breiman [17], in the *scikit-learn* library, the ensemble classifier combination is obtained by averaging over the trees’ probabilistic prediction, instead of the approach in which each tree in the ensemble votes for a single class.

A decision tree ensemble randomization method alternative to random forests was proposed in [16], this method is called extremely randomized trees, which takes randomization one step further, selecting a cut-point at random, independently of the target variable, at each tree node there is also a random choice of a number of attributes among which the best one is determined [16]. The method generates a higher diversity of trees, since it builds randomized trees whose structures are independent of the target variable values of the learning sample. The method shares with random forests the fact that a random subset of candidate features is used for tree expansion, but, instead of looking for the most discriminative thresholds, these thresholds are randomly drawn for each candidate feature, such that the thresholds with the best performance are used for splitting.

This method is aimed not only at increasing accuracy through the threshold randomization, it also reduces the variance of the ensemble since it produces a greater diversity of trees that are randomized in such a way that is independent of the output values of the learning sample [16]. A major difference is that the method does not apply bootstrap samples, using, instead, the whole training dataset. However, one can also implement the method with bootstrapping and OOB scoring, incorporating Breiman’s bagging [17], a feature that is allowed by the *scikit-learn* library and that we will employ and compare with the case without bootstrapping.

The boosting methods that we will test are AdaBoost and gradient boosting methods. AdaBoost predates random forests, indeed, Breiman [17] introduced random forests as an alternative method to AdaBoost, which was developed by Freund and Schapire [20]. AdaBoost’s main approach is to fit an ensemble of weak learners, small trees for instance, repeatedly on modified versions of the training data, the weak learners are aimed at avoiding overfitting, while the repeated use of the training data allows the method to work well on small training datasets.

The boosting iterations results from applying weights to each training sample. For each base learner, the method works with pairs of values for the feature variables and target. In the first step, the initial probability for a training pair to be used for the *j*-th learner is set equal to 1/*N*, now, at each step these probability weights are modified and the learning algorithm is reapplied, in such a way that the training examples that were incorrectly predicted have their weights increased while those that were correctly predicted by the model have their weights decreased, so that the ensemble increasingly learns to better predict that which it failed to predict well in the previous iteration. At the end of training, the ensemble-based prediction is such that the predictions from all of the learners are combined through a weighted majority vote for a final prediction.

Like random forests, the method also reduces variance and increases prediction performance and was introduced as an extension of online prediction to a general decision-theoretical setting [20], which was applied in the original article to a general class of learning problems involving decision making, including gambling, multi-outcome prediction and repeated games, indeed, Freund and Schapire’s proposed method was introduced in the original article as a *decision-theoretic generalization of online learning and an application to boosting* [20].

Even though random forests were proposed by Breiman as an alternative to AdaBoost, the two methods can be combined, indeed, Leshem and Ritov [21] did combine these two methods, using random forests as the base learners, applying random forests boosted with AdaBoost to predict traffic flow, this led to boosted random forests as the next type of boosting algorithm, that combines the randomization methods and bagging with the AdaBoost.

Boosted random forests have been most recently applied to COVID-19 in [19], using patients’ geographical, travel, health and demographic data to predict the severity of the case and the possible outcome, including, recovery or death, with an accuracy of 94% and an F1 Score of 0.86, the analysis developed in [19] revealed that death rates were higher among Wuhan natives when compared to non-natives and that male patients had a higher death rate when compared to female patients, with the majority of affected patients in ages ranging from 20 to 70 years.

We will expand here on boosted random forests and include also, in the tested models for COVID-19 death probability prediction, boosted extremely randomized trees with and without bootstrapping.

Besides standard AdaBoost, boosted random forests and boosted extremely randomized trees, we will also test two other boosting methods: gradient boosting decision trees and histogram gradient boosting. Gradient boosting methods have been successfully applied to COVID-19 prognostic prediction using as features lactic dehydrogenase (LDH), lymphocyte and high-sensitivity C-reactive protein (hs-CRP) [6].

The gradient boosting decision trees method, introduced by Friedman [22,23], combines, sequentially, decision trees as base learners so that each new decision tree fits to the residuals from the previous step, which increases the accuracy and accelerates the learning process.

Recently, in [24], a new gradient boosting method called LightGBM was proposed, which, as the authors showed, was capable of outperforming leading gradient boosting decision trees’ accelerating algorithms, including XGBoost, both in terms of training time and accuracy. In this sense, LightGBM is a big data scalable method with state-of-the-art results. The method has been applied to EEG analysis and has been shown to have state-of-the-art performance when compared with convolutional neural networks, gated recurrent units, support vector machines, and large margin nearest neighbor models, outperforming these models in real-time EEG mental state prediction [25].

The LightGBM-based histogram gradient boosting algorithm, which is implemented by *scikit-learn*, bins the training data, which may be an advantage for quantitative continuous-valued feature variables, however, it may also be seen as a disadvantage for discrete qualitative data, and one needs to be careful in considering the ordering of variables in a numeric recoding process, however, if feature variables are ordinal, and a good recoding is achieved, defining, for binary variables, the order in terms of the Bernoulli success, with one-hot encoding used for qualitative nominal variables, this method can still be used to accelerate the gradient boosting learning process, where the bins can lead to the discovery of patterns that may increase accuracy, especially in the case of many-features leading to a combinatorially sufficiently high number of alternative values for the full discrete finite feature space.

For our main dataset, the binning done by *sciki-learn*’s LightGBM-based method becomes equivalent to defining subsets from the whole discrete feature space, which is a finite set of size 36, accelerating the gradient boosting process, the maximum number of bins can, therefore, in our case, be set to 36.

Considering the above review, we will be comparing the following major tree-based ensemble machine learning models in COVID-19 death probability prediction on the CDC dataset:

- Random forest (RF);
- Extremely randomized trees (ERTs);
- Extremely randomized trees with bootstrap sampling and OOB scoring (BERTs);
- AdaBoost;
- Boosted Random Forest (Boosted RF);
- Boosted extremely randomized trees (Boosted ERTs);
- Boosted extremely randomized trees with bootstrap sampling and OOB scoring (Boosted BERTs);
- Gradient Boosting Trees (GBTs);
- Histogram Gradient Boosting (HGB).

We also compare the performance of the above methods with that of logistic regression, used as a basic benchmark, we chose the logistic regression for a benchmark, since it is a major machine learning model used when dealing with probability prediction in risk contexts, especially in the healthcare context, and it has been successfully applied to COVID-19 symptoms modeling [26]. For performance evaluation, we use the metrics reviewed in the previous subsection and choose the best performer on test data to extract the probability profile for further analysis.

## 3. Results

### 3.1. Main Results and Performance Evaluation

In this section, we analyze the performance of the logistic regression and the main tree-based ensemble models reviewed in the previous section. The training set was a randomly chosen sample of 413,800 cases, leaving 413,801 cases in the test sample, which gives an around 50/50 distribution of cases.

For the logistic regression, the inverse of regularization strength was set equal to 10, we also employed 5-fold cross-validation with L2 penalty and lbfgs solver. For the RF, ERTs, boosted RF and boosted ERTs models, we used 100 trees, and Gini impurity measure.

In the case of the RF model, we employed bootstrap sampling with OOB scoring, in the case of the ERTs, we employed the method without bootstrap sampling and with bootstrap sampling which also employed OOB scoring.

For the boosting algorithms, the AdaBoost used 100 trees of maximum depth equal to 2, and SAMME.R algorithm, the maximum number of features when looking for the best split was set to the square root of the number of features^5^. In the case of the boosted RFs and boosted ERTs we also used 100 base estimators, each comprised of 100 trees.

For the GBTs we used Friedman mean squared error, 100 estimators and a subsample^6^ of 75%, for the HGB we used a learning rate of 7% and an L2 regularization of 2%, the maximum number of bins was set equal to 36 which is the full size of the discrete feature space, we also did not employ early stopping^7^.

In **figure 3**, we show the logistic regression’s reliability plots, for the training and test data, while, in the upper plot region, the predicted probability versus the observed proportion shows a good fit, there is some dispersion in the lower region.

**Fig. 3:**
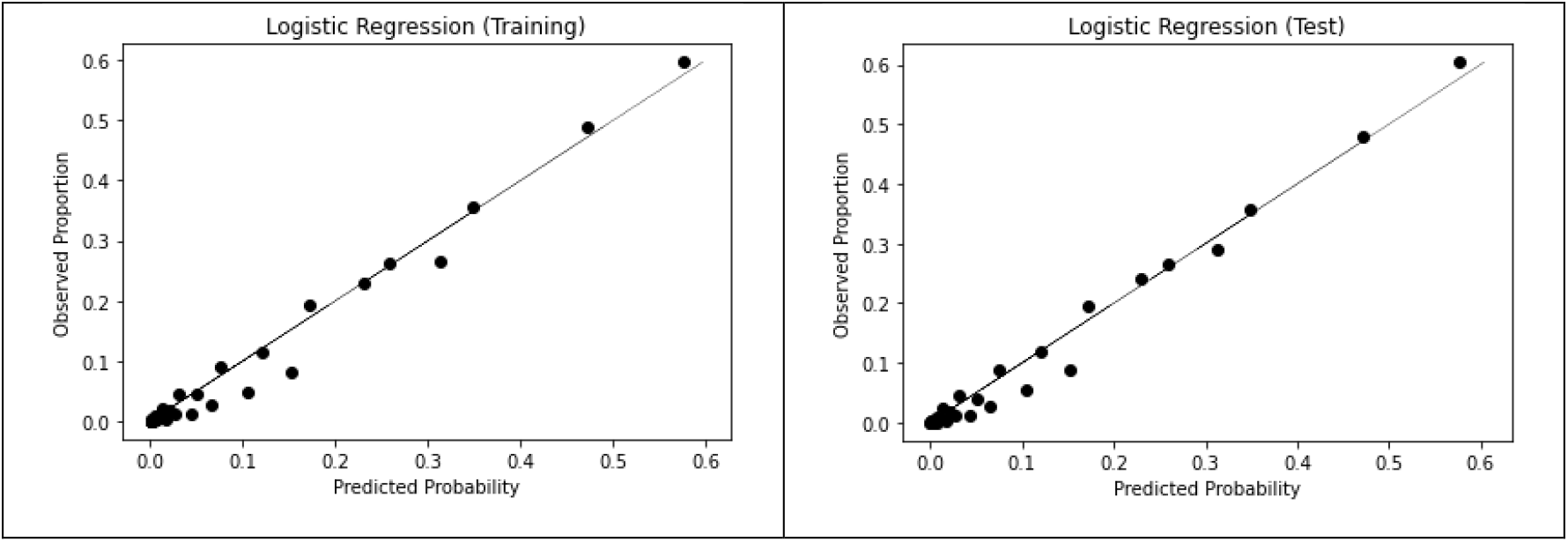
Training and test data reliability plots for the logistic regression.

Considering the RFs, ERTs and BERTs, as shown in **figure 4**, all models exhibit a good fit to the diagonal line in the reliability plots, therefore, in terms of visual analysis, all these models seem to show a good calibration, that also seems to be better than the logistic regression model.

**Fig. 4:**
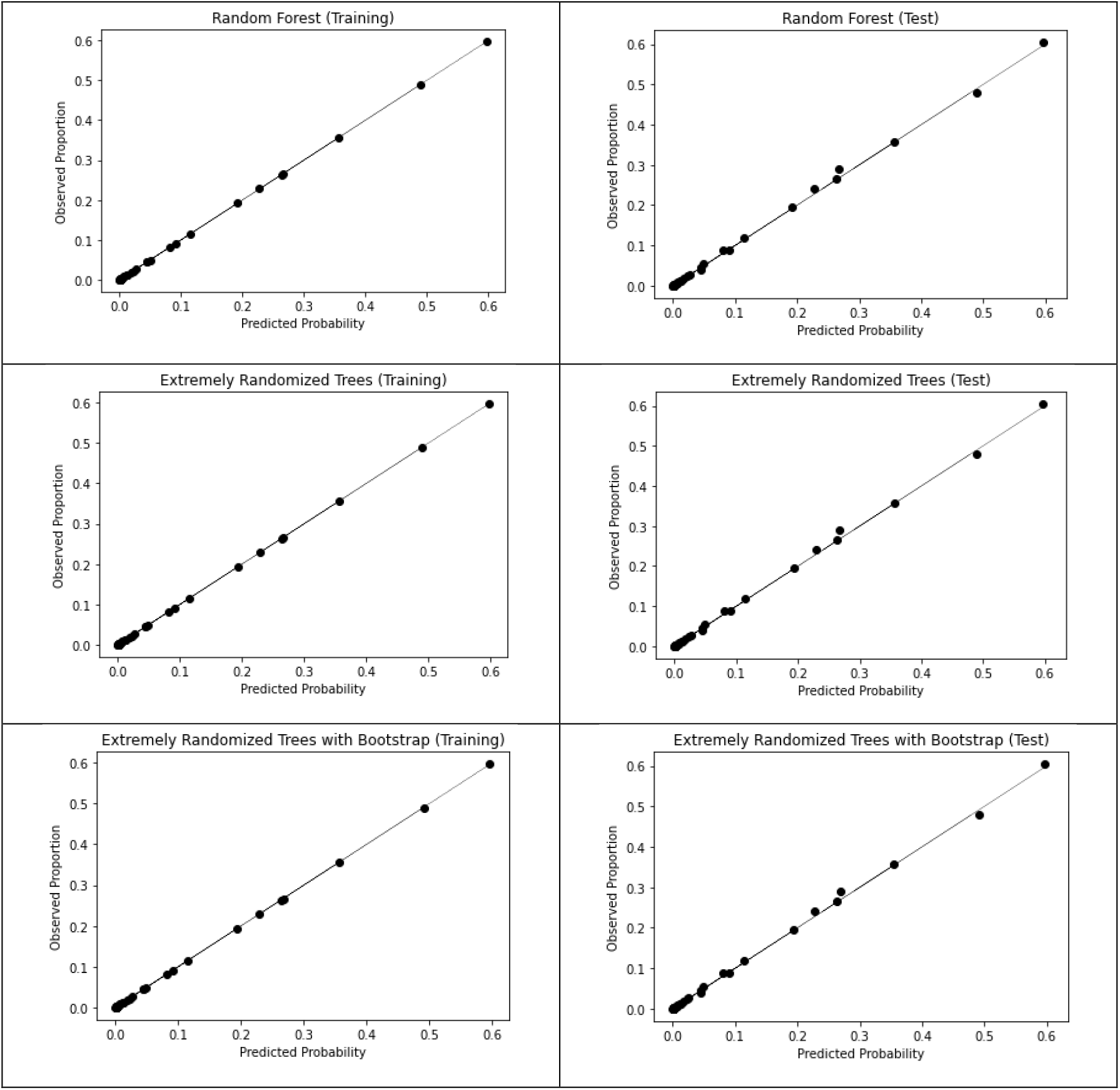
Training and test data reliability plots for the RF, ERTs and BERTs.

Evaluating, now, the reliability plots for AdaBoost, boosted RFs, boosted ERTs and boosted BERTs, unlike in the previous models, we find a poor calibration, as can be seen in the reliability plots shown in **figure 5**.

**Fig. 5:**
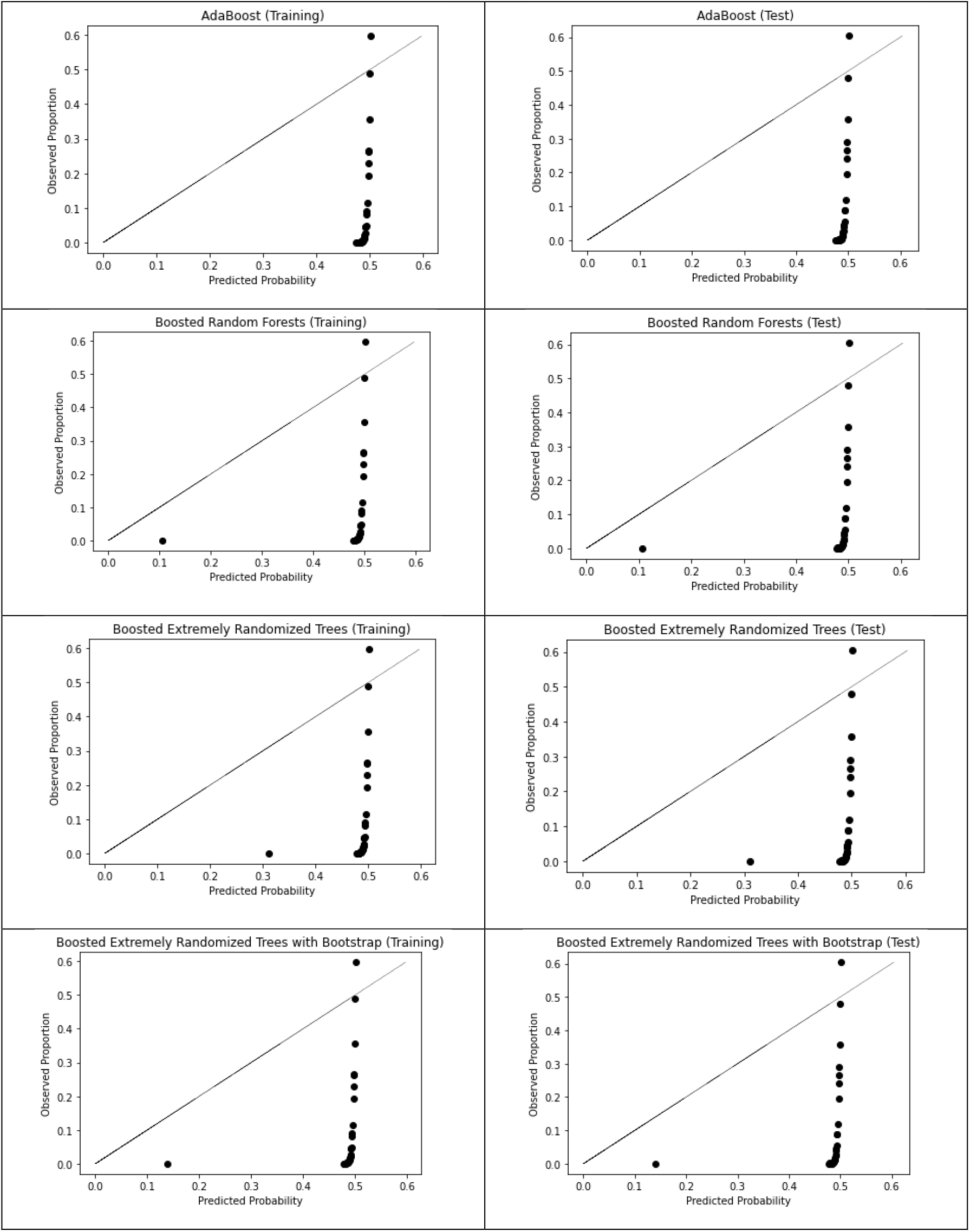
Training and test data reliability plots for the AdaBoost with decision trees, boosted RF, boosted ERTs and boosted BERTs.

In order to work these models, we need to employ calibration methods, in order to correct for the poor calibration [11].

Platt’s calibration and isotonic regression, which is a generalization of Platt’s method by fitting an isotonic function instead of fitting a sigmoid as in Platt’s calibration, are the two main methods of calibration that have been worked for binary targets [11] and that can be used to correct for miscalibration.

For this dataset, we found a better performance using isotonic calibration, so we applied isotonic calibration to the four boosted models that employed AdaBoost.

After calibration, all the four models exhibit a good fit to the perfect calibration line (the main diagonal) in the reliability plot, as shown in **figure 6**.

**Fig. 6:**
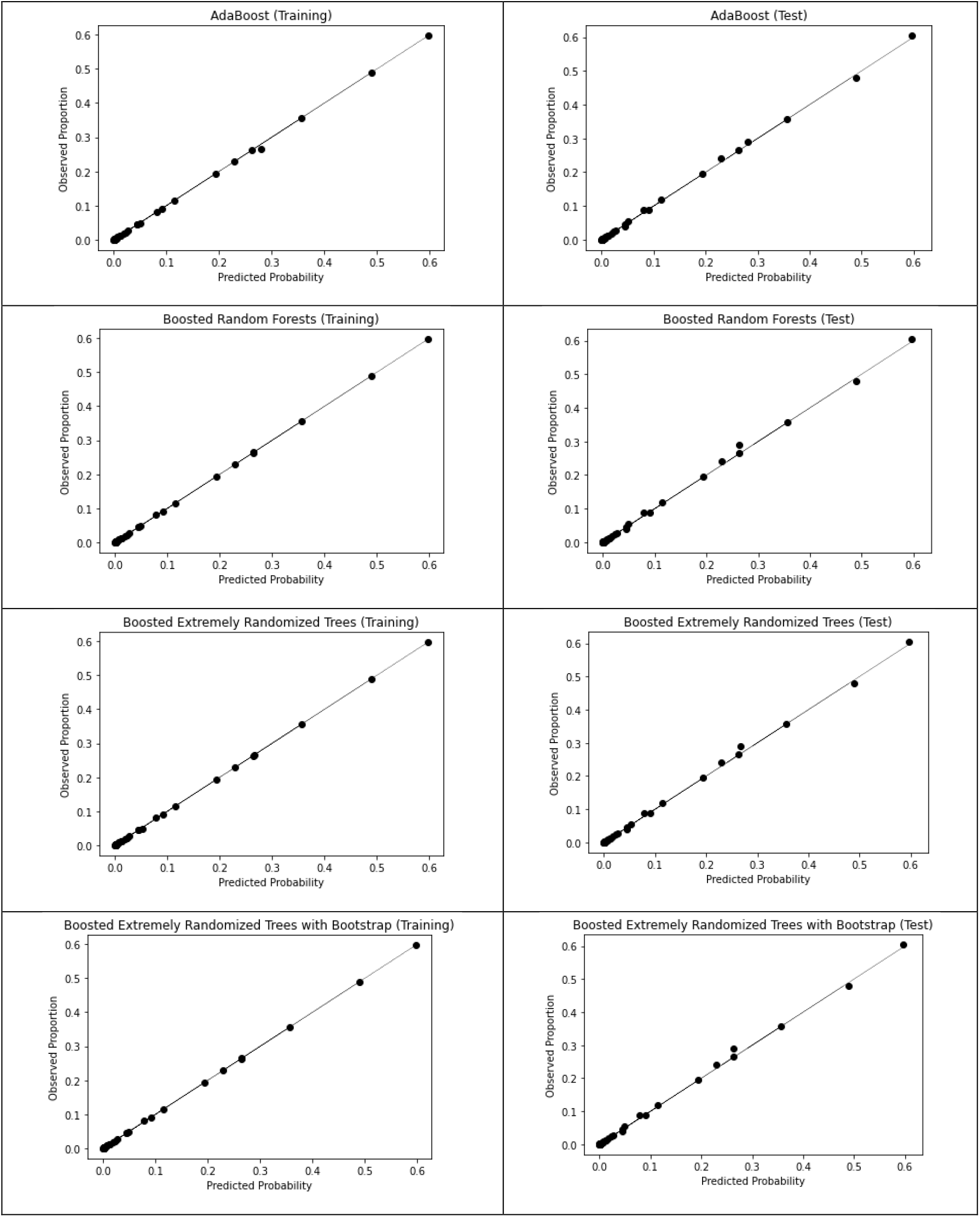
Training and test data reliability plots for the calibrated AdaBoost, calibrated boosted RF, calibrated boosted ERTs and calibrated boosted BERTs.

The two other gradient boosting models GBTs and HGB show a good fit both in training and in testing, without the need for the application of a calibration method, as shown in **figure 7**.

**Fig. 7:**
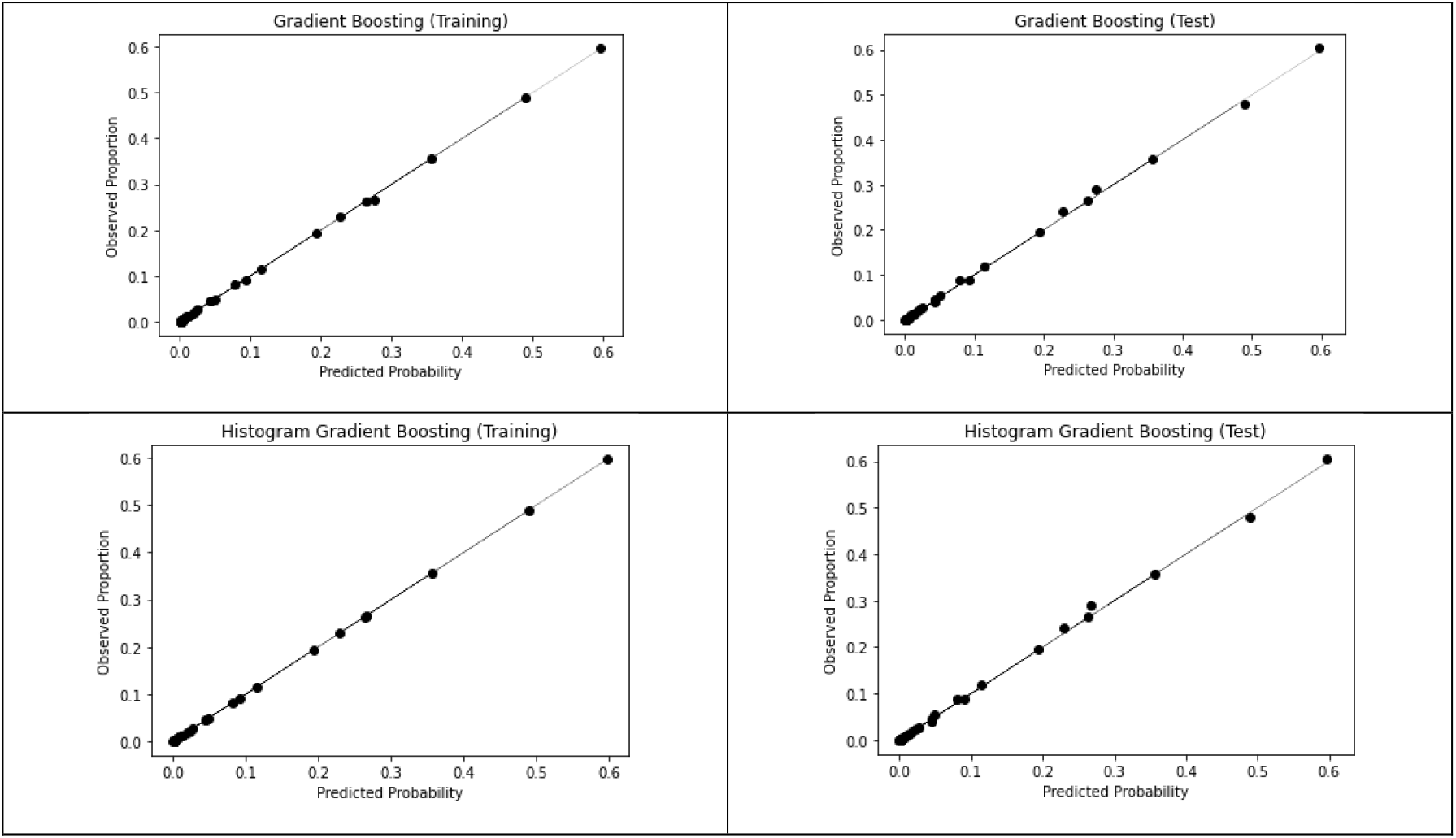
Training and test data reliability plots for GBTs and HGB.

After analyzing the main reliability plots, we now need to analyze the main calibration performance metrics which are shown in tables 3 and 4. Considering the results shown in these tables, we find that all machine learning models exhibit a high sample calibration performance, in both training and testing, with all models having a more than 98% of reliability plot explained variance.

**Table 3:**
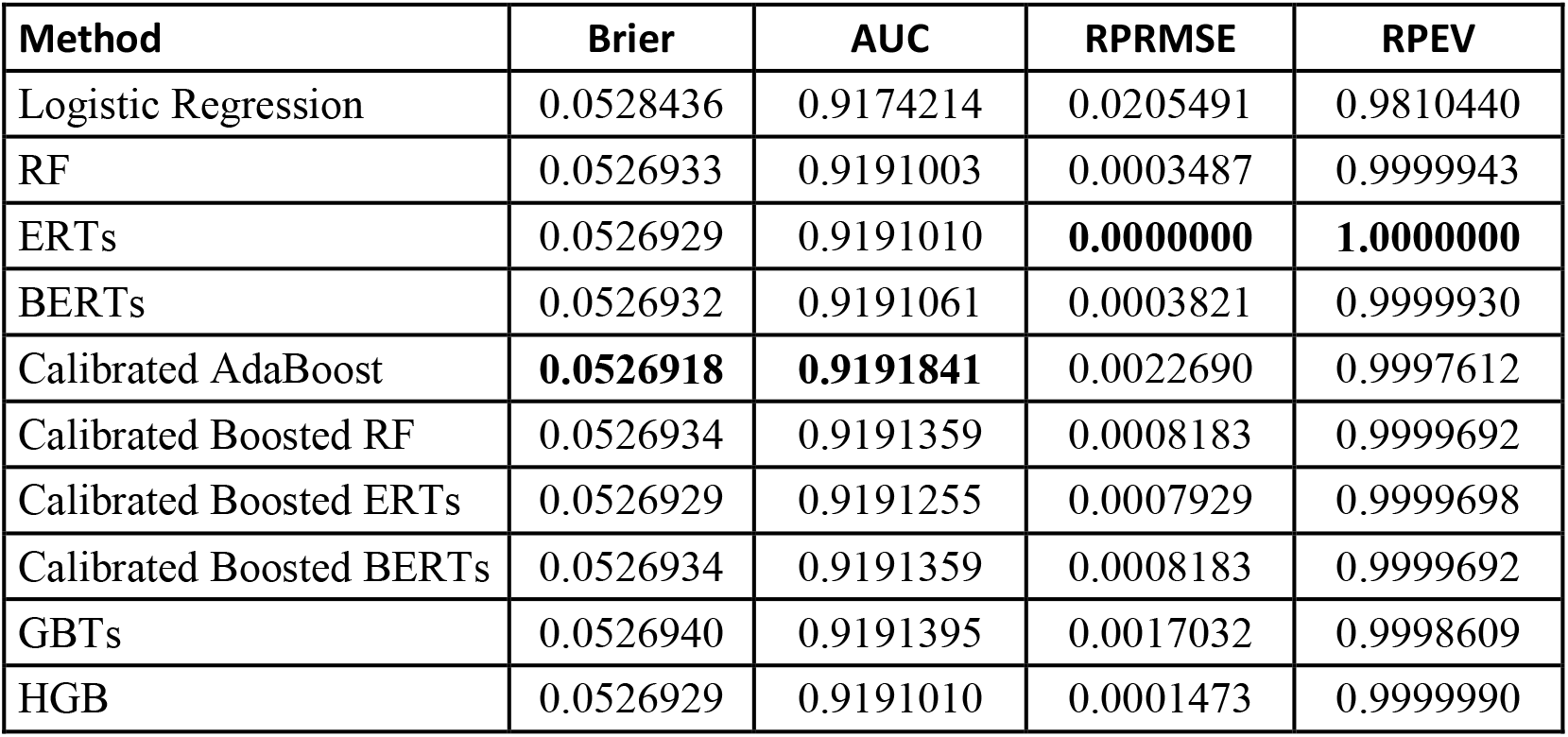
Main metrics for the machine learning models in the training data.

| Method | Brier | AUC | RPRMSE | RPEV |
| --- | --- | --- | --- | --- |
| Logistic Regression | 0.0528436 | 0.9174214 | 0.0205491 | 0.9810440 |
| RF | 0.0526933 | 0.9191003 | 0.0003487 | 0.9999943 |
| ERTs | 0.0526929 | 0.9191010 | **0.0000000** | **1.0000000** |
| BERTs | 0.0526932 | 0.9191061 | 0.0003821 | 0.9999930 |
| Calibrated AdaBoost | **0.0526918** | **0.9191841** | 0.0022690 | 0.9997612 |
| Calibrated Boosted RF | 0.0526934 | 0.9191359 | 0.0008183 | 0.9999692 |
| Calibrated Boosted ERTs | 0.0526929 | 0.9191255 | 0.0007929 | 0.9999698 |
| Calibrated Boosted BERTs | 0.0526934 | 0.9191359 | 0.0008183 | 0.9999692 |
| GBTs | 0.0526940 | 0.9191395 | 0.0017032 | 0.9998609 |
| HGB | 0.0526929 | 0.9191010 | 0.0001473 | 0.9999990 |

**Table 4:** Main metrics for the machine learning models in the test data.

| Method | Brier | AUC | RPRMSE | RPEV |
| --- | --- | --- | --- | --- |
| Logistic Regression | 0.0527035 | 0.9179402 | 0.0184920 | 0.9844724 |
| RF | 0.0525347 | 0.9196267 | 0.0056403 | 0.9986259 |
| ERTs | **0.0525347** | **0.9196268** | 0.0054633 | 0.9987093 |
| BERTs | 0.0525348 | 0.9196267 | 0.0053414 | 0.9987733 |
| Calibrated AdaBoost | 0.0525351 | 0.9196238 | **0.0039613** | **0.9993227** |
| Calibrated Boosted RF | 0.0525348 | 0.9196226 | 0.0059652 | 0.9984652 |
| Calibrated Boosted ERTs | 0.0525348 | 0.9196262 | 0.0054924 | 0.9986929 |
| Calibrated Boosted BERTs | 0.0525348 | 0.9196226 | 0.0059652 | 0.9984652 |
| GBTs | 0.0525355 | 0.9196060 | 0.0049337 | 0.9989557 |
| HGB | 0.0525347 | **0.9196268** | 0.0055371 | 0.9986714 |

The logistic regression has a good performance, with a Brier loss, for the training data, of 0.0528436 and, for the test data, of 0.0527035. The AUC is of 0.9174214, for the training data, and of 0.9179402, for the test data.

Considering the reliability plot’s RMSE and explained variance, the logistic regression has a training data RPMSE of 0.0205491 and a test data RPMSE of 0.0184920, the training data RPEV is 0.9810440 and the test data RPEV is 0.9844724, which shows that the logistic regression’s COVID-19 predicted death probabilities, conditional on the feature values, explains slightly more than 98% of the variability of the conditional sample proportions.

Overall these results show a good performance of the logistic regression in COVID-19 death probability prediction, however, when we turn to the ensemble tree-based models, we find that they all perform even better than the logistic regression, with a RPEV that is higher than 0.99, in both training and test data, that is, these models’ probability predictions, conditional on the feature values, account for more than 99% of the conditional sample proportions’ variability. The RPRMSE for all tree-based models also show a value lower than 1%, both in the training data and in the test data, outperforming the logistic regression in the reliability plot metrics, which confirms our previous visual analysis.

Considering the Brier loss, we find that the best performer in the training data is the AdaBoost with isotonic calibration, which is also the model with the highest AUC, in the test data, the best performer, in accordance with the Brier loss, is the ERTs model^8^, which, along with the HGB model, has the highest AUC^9^.

However, the Brier loss can be misleading, as argued in [13]. If we look at the RPRMSE and RPEV we find an inverse relation: the ERTs model is the best performer in training but not in the test data, where we find that the best performer is the isotonic calibrated AdaBoost.

For the ERTs model, there seems to be an overfitting in training, where the ERTs capture the training sample proportions with approximately 100% explained variance and 0 RPRMSE, but when we look at the test data, the RPRMSE rises to 0.0054633 and the RPEV decreases to 0.9987093. The best performer in test data is the calibrated AdaBoost which is the only one with higher than 0.999 test data RPEV, with all the other tree-based algorithms exhibiting RPEV slightly higher than 0.998 but lower than 0.999, in the test data. Also, with the exception of the GBTs and calibrated AdaBoost models, all the ensemble tree-based models have a test data RPEV that is higher than 0.005.

The calibrated AdaBoost model does not fit as well as the others in the training data, in what regards the reliability plot metrics, but it gains in generalizability with relatively stable results in these metrics, from training to testing, showing an RPRMSE of 0.0022690 in the training data and of 0.0039613 in the test data, which shows that there is no significant loss of calibration performance, a similar pattern can be seen in the RPEV which has a value of 0.9997612 in the training data and of 0.9993227 in the test data, showing a consistent performance in both training and test data.

Given these results, we will use the calibrated AdaBoost model for the profiling. Nevertheless, we show, in the appendix, the death probability profile provided by the calibrated AdaBoost side by side with the one provided by the ERTs, the deviation between the two algorithms is not that significant, with the largest deviation being of around 0.013394 (which occurs for male individuals with no underlying comorbidity or disease of the 80+ age group), from table 5’s results, shown in the appendix, it follows that the general pattern of the profile that we now analyze for the calibrated AdaBoost also holds for the ERTs, a point that reinforces the robustness of the findings, it is also worth stressing that the test data Brier score difference between the calibrated AdaBoost and the ERTs is small, of around 4.434E-07.

**Table 5:** Probability profiles for the Calibrated AdaBoost and the ERTs.

| Sex | Age Group | Medical Conditon | Calibrated AdaBoost | ERTs | Deviation |
| --- | --- | --- | --- | --- | --- |
| Female | 0 - 9 Years | No | 0.000662 | 0.000672 | 9.847985E-06 |
| Female | 0 - 9 Years | Yes | 0.001094 | 0.000814 | 2.807287E-04 |
| Female | 10 - 19 Years | No | 0 | 0 | 0.000000E+00 |
| Female | 10 - 19 Years | Yes | 0.001541 | 0.001801 | 2.594068E-04 |
| Female | 20 - 29 Years | No | 0.000194 | 0.000216 | 2.281746E-05 |
| Female | 20 - 29 Years | Yes | 0.002962 | 0.003243 | 2.815308E-04 |
| Female | 30 - 39 Years | No | 0.000662 | 0.000734 | 7.190387E-05 |
| Female | 30 - 39 Years | Yes | 0.007570 | 0.007668 | 9.805658E-05 |
| Female | 40 - 49 Years | No | 0.001543 | 0.001790 | 2.469486E-04 |
| Female | 40 - 49 Years | Yes | 0.018027 | 0.017878 | 1.488485E-04 |
| Female | 50 - 59 Years | No | 0.004398 | 0.003963 | 4.350348E-04 |
| Female | 50 - 59 Years | Yes | 0.044294 | 0.044567 | 2.725841E-04 |
| Female | 60 - 69 Years | No | 0.012529 | 0.012203 | 3.259155E-04 |
| Female | 60 - 69 Years | Yes | 0.114547 | 0.114555 | 7.870341E-06 |
| Female | 70 - 79 Years | No | 0.050531 | 0.048760 | 1.770816E-03 |
| Female | 70 - 79 Years | Yes | 0.262284 | 0.263278 | 9.941172E-04 |
| Female | 80+ Years | No | 0.229577 | 0.228674 | 9.027776E-04 |
| Female | 80+ Years | Yes | 0.490248 | 0.490292 | 4.368747E-05 |
| Male | 0 - 9 Years | No | 0.000520 | 0.000435 | 8.496840E-05 |
| Male | 0 - 9 Years | Yes | 0.000662 | 0.000713 | 5.107174E-05 |
| Male | 10 - 19 Years | No | 0.000122 | 0.000154 | 3.215310E-05 |
| Male | 10 - 19 Years | Yes | 0.002263 | 0.002217 | 4.581467E-05 |
| Male | 20 - 29 Years | No | 0.000520 | 0.000406 | 1.143676E-04 |
| Male | 20 - 29 Years | Yes | 0.007570 | 0.007817 | 2.468476E-04 |
| Male | 30 - 39 Years | No | 0.001541 | 0.001342 | 1.994819E-04 |
| Male | 30 - 39 Years | Yes | 0.022310 | 0.022233 | 7.675542E-05 |
| Male | 40 - 49 Years | No | 0.004846 | 0.004935 | 8.865559E-05 |
| Male | 40 - 49 Years | Yes | 0.044693 | 0.044556 | 1.370955E-04 |
| Male | 50 - 59 Years | No | 0.010987 | 0.011164 | 1.774924E-04 |
| Male | 50 - 59 Years | Yes | 0.090999 | 0.090938 | 6.053668E-05 |
| Male | 60 - 69 Years | No | 0.026159 | 0.026145 | 1.400498E-05 |
| Male | 60 - 69 Years | Yes | 0.193004 | 0.192963 | 4.112802E-05 |
| Male | 70 - 79 Years | No | 0.081416 | 0.081068 | 3.483084E-04 |
| Male | 70 - 79 Years | Yes | 0.355986 | 0.355941 | 4.483705E-05 |
| Male | 80+ Years | No | 0.280218 | 0.266824 | 1.339354E-02 |
| Male | 80+ Years | Yes | 0.596808 | 0.596867 | 5.922762E-05 |

### 3.2. COVID-19 Death Probability Profile

In **figure 8** and table 6, we show the probability profile produced by the calibrated AdaBoost model, we find that the predicted death probability increases exponentially with age, a pattern that we also saw in the inferential analysis for the age factor, but the rate of this increase depends upon the sex and medical condition.

**Table 6:** Probability profile for the age group, sex and underlying comorbidity or disease, extracted from the calibrated AdaBoost model.

| Age Group | No Underlying Comorbidity or Disease |  | With Underlying Comorbidity or Disease |  |
| --- | --- | --- | --- | --- |
|  | Male | Female | Male | Female |
| 0 - 9 Years | 0.000520 | 0.000662 | 0.000662 | 0.001094 |
| 10 - 19 Years | 0.000122 | 0.000000 | 0.002263 | 0.001541 |
| 20 - 29 Years | 0.000520 | 0.000194 | 0.007570 | 0.002962 |
| 30 - 39 Years | 0.001541 | 0.000662 | 0.022310 | 0.007570 |
| 40 - 49 Years | 0.004846 | 0.001543 | 0.044693 | 0.018027 |
| 50 - 59 Years | 0.010987 | 0.004398 | 0.090999 | 0.044294 |
| 60 - 69 Years | 0.026159 | 0.012529 | 0.193004 | 0.114547 |
| 70 - 79 Years | 0.081416 | 0.050531 | 0.355986 | 0.262284 |
| 80+ Years | 0.280218 | 0.229577 | 0.596808 | 0.490248 |

**Fig. 8:**
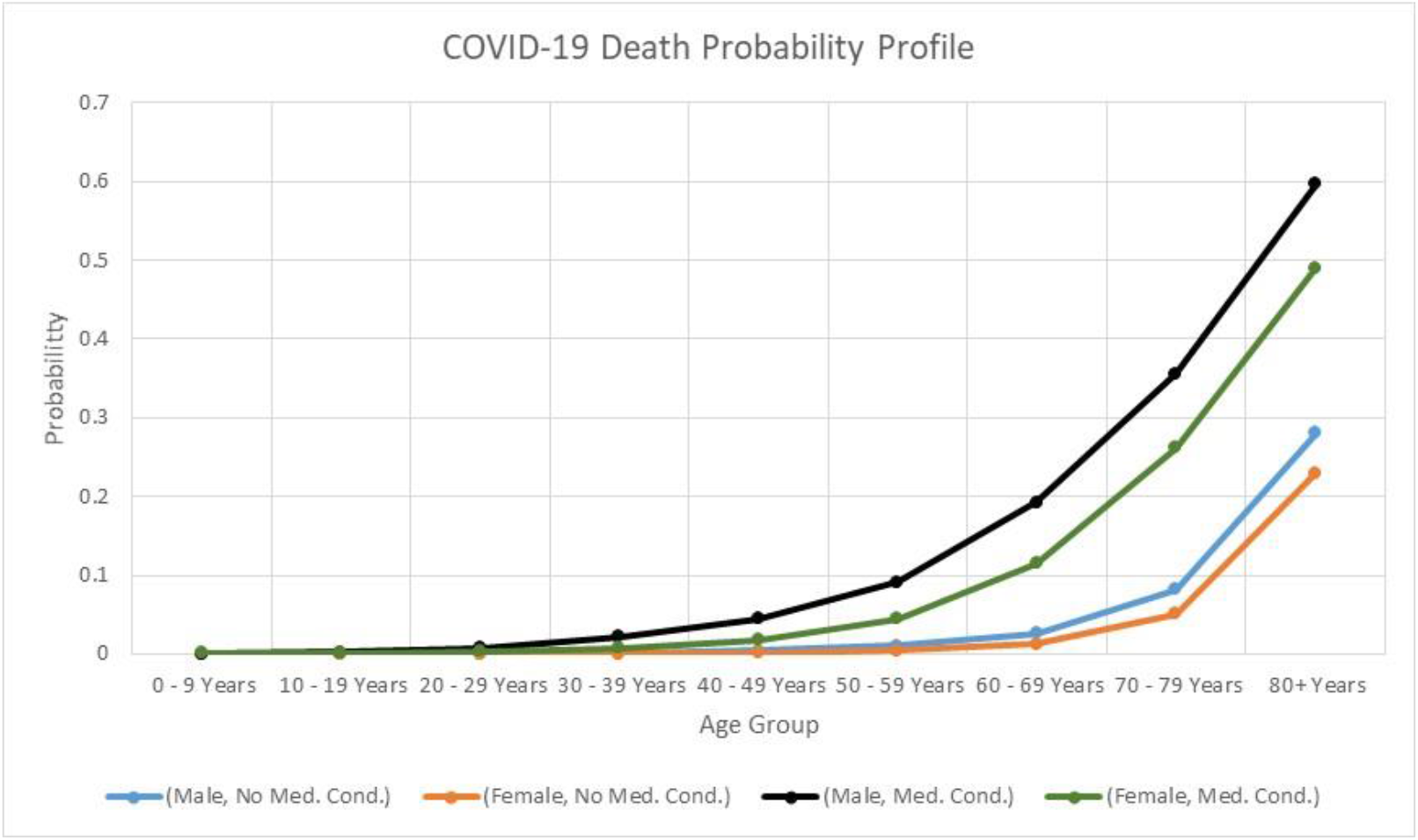
Probability profile plot for the age group, sex and underlying comorbidity or disease, extracted from the calibrated AdaBoost model.

The first point that one can notice is that age and the medical condition variables seem to critically affect the death probability, with the male sex being a factor in increasing the death risk. Furthermore, the risk difference between male and female sex seems to increase with the age group, so that while both males and females have an exponentially increasing death probability, the males show a more rapid exponential rise in the death probability than females. These differences between males and females become more significant for people with underlying comorbidity or disease.

Considering people with no underlying comorbidity or disease, according to the isotonic calibrated AdaBoost model, the 1% death probability is surpassed at the age group 50-59 in males and at 60-69 in females, for this last age group, the calibrated AdaBoost model assigns to males a death probability higher than 2%. At 70-79 the males have an assigned death probability slightly higher than 8%, while the females have an assigned death probability slightly higher than 5%, however, at 80+ both males and females, with no underlying comorbidity or disease, have a higher than 20% assigned death probability, with the males having a death probability, assigned by the model, slightly higher than 28% and the females near 23%. This is the highest difference between the two sexes for people with no underlying comorbidity or disease, which is an around 5% difference.

If we compare this difference with that obtained for the same age group, 80+, but with underlying comorbidity or disease, we find a difference near 11% between the two sexes, which shows that, while being of the male sex is a differentiating risk factor, it is a much higher risk factor for people with underlying comorbidity or disease.

Overall, the general profile for people with no underlying comorbidity or disease is close for both sexes, with the more critical age groups being situated in the ages of 70-79 and 80+ years.

In the case of people with underlying comorbidity or disease, the critical threshold for high risk group is much lower, indeed, at 50-59 years, the males with underlying comorbidity or disease have an assigned death probability by the model that is already higher than the 70-79 males with no underlying comorbidity or disease, which indicates that 50-59 males with underlying comorbidity or disease can be considered as a group with comparable risk to that of 70-79 males with no underlying comorbidity or disease.

If we consider the 1% death probability mark, we find that while this threshold, for individuals with no underlying comorbidity or disease, is surpassed at the age group of 50-59 years for males and at 60-69 years for females, when we consider individuals with underlying comorbidity or disease, the 1% death probability threshold is surpassed at age group 30-39 for males and at 40-49 for females. The 1% death probability threshold is thus surpassed with an around 20 years reduction in the age group, for people with underlying comorbidity or disease in comparison with those that do not have an underlying comorbidity or disease.

For the age group 60-69, the model assigns a death probability for males with underlying comorbidity or disease that is near 20%, at 70-79 this probability rises to near 35.6% and at 80+ years it is near 60%. For the females we find a slower build up with a growing distance from the males, but, even so, we have a high risk for females with underlying comorbidity or disease from the 50-59 age group onwards, with a more than 11% assigned death probability for the 60-69 age group, which is higher than the females with no underlying comorbidity or disease of the 70-79 age group, for this last group the females with underlying comorbidity or disease have an assigned death probability slightly higher than 26% and the 80+ group shows a rise to around 49%. Having analyzed the main profile, we now discuss these results.

## 4. Discussion

Our results show that machine learning models can be applied to successfully capture the COVID-19 death probability profiles, conditional on three main identified risk factor variables: sex, age and underlying comorbidity or disease. The tree-based ensemble machine learning models that we compared, when applied to the qualitative variables that comprise the CDC’s database for these factors, were capable of capturing more than 99% explained variance of the reliability plots, in both training and test data, showing an improved performance over the logistic regression model which captured 98.10440% of the training data reliability plot’s explained variance and 98.44724% of the test data reliability plot’s explained variance. While the AdaBoost-based models (AdaBoost, boosted RF, boosted ERTs and boosted BERTs) all showed poor calibration, after isotonic calibration, all exhibited a good calibration and the isotonic calibrated AdaBoost was the best performing model in the test data reliability plot-based metrics, both in regards to the reliability plot’s RMSE (0.0039613) and explained variance (0.9993227), in the training data these metrics were of 0.0022690 and 0.9997612, respectively.

The calibrated AdaBoost also showed the lowest Brier loss in the training data, though in the test data the lowest Brier loss was obtained by the ERTs model, however, the difference between the two models’ test data Brier loss is of around 4.434E-07. Given the reliability plot performance of the calibrated AdaBoost we analyzed that model’s results, even though the overall profiles produced by these two models are close to each other, so that the main findings do not change.

The resulting profile confirms and reinforces previous studies’ results [1-5, 18, 26, 27] on COVID-19 risk factors. The profile is also consistent with the exponential increase in death risk with age, that we analyzed inferentially. The results also show that the definition of what is a risk group needs to factor age, sex and underlying comorbidity or disease with segmented risk group profiling, indeed, males are in general more at risk, but their risk gap with respect to female patients increases when underlying comorbidity or disease is present.

In the COVID-19 death risk profile, while the presence of an underlying comorbidity or disease lowers the high risk age threshold and operates as an exponential accelerator of death probability, by increasing the exponential growth rate of death probability with age, it also rises significantly the death probability values for the high risk age groups, explaining the reason for the death proportion’s 95% confidence intervals that we calculated in the exploratory data analysis for the age groups.

In this sense, from the machine learning profile, it follows that the 95% confidence interval for the 70-79 years age group that situates, on an inference level, the population’s death proportion between 0.272681 and 0.279785, and the 95% confidence interval for the 80+ years age group, that situates the population’s death proportion between 0.505483 and 0.513633, may be largely driven by the combined effect of age and underlying comorbidity or disease, which leads, in the case of the age group 70-79, for individuals with underlying comorbidity or disease to a predicted death probability of 0.355986 for males and 0.262284 for females, likewise, in the case of the 80+ age group, these probabilities rise to 0.596808 in the case of males and to 0.490248 in the case of females.

The fact that males with no underlying comorbidity or disease also tend to have higher death risk than females as age progresses also reinforces the findings of an asymmetric impact of COVID-19 in regards to sex and is consistent with previous studies’ results [1-5]. A possible explanation provided in [5], regarding male infectiousness, relates to smoking and higher circulating angiotensin converting enzyme 2 (ACE2) levels in men, SARS-CoV-2 utilizes ACE2 receptors found at the surface of the host cells to get inside the cell, this is also an important factor in specific underlying comorbidities or diseases.

As researched in [27], patients with obesity, diabetes, chronic obstructive pulmonary disease, cardiovascular diseases, hypertension, malignancies, HIV and other underlying comorbidities or diseases develop life-threatening situations. Underlying comorbidities or diseases that are associated with a strong ACE2 receptor expression can, as argued in [27], enhance the viral entry of SARS-CoV-2 into host cells, this factor along with weakened immune systems may work as accelerators of life threatening conditions and may help explain the exponential increase in death probability with age, an exponential increase that shows, in the probability profile produced by the calibrated AdaBoost model, an accelerated exponential growth rate when an underlying comorbidity or disease is present^10^, as we saw above.

Common comorbidities that show high COVID-19 fatality rates, include obesity, liver diseases, renal diseases, chronic obstructive pulmonary disease, cardiovascular diseases, diabetes, hypertension and malignancy standing out, with fatality rates, as reviewed in [27], respectively of 68% (for obesity), 29% (for liver diseases), 26% (for renal diseases), 20% (for chronic obstructive pulmonary disease), 15% (for cardiovascular diseases), 8% (for diabetes), 6% (for hypertension) and 2% (for malignancy).

Returning to the model’s results, if, in terms of health policy, one fixes age-based high risk groups at 70-79 onwards, or even at 60-69 onwards, this can lead to a flawed representation of the true death risk, the machine learning model indicates that we need to set the high risk age groups differentially on sex and underlying comorbidity or disease, indeed, we found the need for lowering the high risk age groups based on sex and underlying comorbidity or disease, thus, for instance, in males with underlying comorbidity or disease, the high risk group can be argued to start at 50-59 years onwards. Indeed, 50-59 and 60-69 males with underlying comorbidity or disease are found by the machine learning model to be of a higher death risk than the 70-79 males with no underlying comorbidity or disease.

These results have an important consequence, since individuals at age 50-59 and 60-69 are professionally active individuals, these individuals are potentially less able to remain confined and may be more exposed, if careful measures are not taken in the workspace, so that while we may expect, with the spread of the disease, not only higher frequencies of hospitalizations of individuals with no underlying comorbidity or disease of the 70-79 and 80+ age groups, which are high risk groups, we may also expect to see rising hospitalizations and possibly intensive care unit pressure coming from individuals of lower age groups, with underlying comorbidity or disease, especially males of the 50-59 age group onwards. For females with underlying comorbidity or disease we can also expect these rising hospitalizations and possibly intensive care unit pressure from the 60-69 age group onwards.

It is also worth stressing that the model identified another relevant point, the 40-49 age group is already of considerable risk for males with underlying comorbidity or disease and with a comparable risk to that of females with underlying comorbidity or disease but of the 50-59 age group.

These results stress the need for further research on the combined impact of age, sex and underlying comorbidity or disease, towards helping explain and predict how different underlying comorbidity or diseases may impact asymmetrically sex and how they can be expressed exponentially with age enhancing the COVID-19 death risk.

## Data Availability

The data used for running and testing the machine learning algorithms was CDC's COVID-19 case surveillance data made publicly available at https://data.cdc.gov/Case-Surveillance/COVID-19-Case-Surveillance-Public-Use-Data/vbim-akqf.

## Footnotes

1 The data is made available for public use at https://data.cdc.gov/Case-Surveillance/COVID-19-Case-Surveillance-Public-Use-Data/vbim-akqf, the version that we use was updated in November 4, 2020.

2 Classified in the database as a Yes or No binary feature.

3 The reverse labeling would correspond to a survival probability, which is an alternative way to approach the problem. If one focuses on death probability, however, the labeling requires that the Bernoulli success case be labeled as 1-dead and 0-alive.

4 Noticeably, while age is a continuous quantitative variable, the CDC’s age group is already pre-binned into nine age groups, becoming an ordinal feature variable, so that for our base dataset no additional binning considerations are involved in obtaining the reliability plot.

5 We tried with other parameters and got worse results for the algorithm.

6 The choice of a subsample was aimed at variance reduction, and was set to value 75% rather than lower due to a tradeoff with bias.

7 We experimented on different parameters and found the results to be resilient for all the models being tested. For the histogram boosting, however, we found that there was a small region of learning rates and regularization parameters that yielded a good performance, a learning rate of 7% and a regularization of 2% yielded the best results.

8 Although the test data Brier score has the same value, up to a seventh decimal place approximation, to that of HGB and RF, the values are actually different. The ERTs model has a Brier score of 0.0525346880585105, while the HGB and the RF models have test data Brier scores of 0.0525347010331709 and 0.0525347253515783, respectively, so that the ERTs model is indeed, the one with the lowest Brier score in the test data.

9 In this case, the two estimated AUC values do coincide.

10 A point that holds for both the isotonic calibrated AdaBoost and the ERTs.

